# DNA methylation patterns associated with cyanogenic cassava exposure and konzo in Sub-Saharan Africa

**DOI:** 10.1101/2021.07.28.21261215

**Authors:** Kristen Kocher, Surajit Bhattacharya, Matthew S. Bramble, Daniel Okitundu-Luwa, Dieudonne Mumba Ngoyi, Desire Tshala-Katumbay, Eric Vilain

## Abstract

**Main Text:** Konzo, a disease characterized by sudden, irreversible spastic paraparesis, affecting up to 10% of the population in some regions of Sub-Saharan Africa during outbreaks and is strongly associated with dietary exposure to cyanogenic bitter cassava. The molecular mechanisms underlying the development of konzo, remain largely unknown. Here, through an analysis of 16 individuals with konzo and matched healthy controls from the same outbreak zones, we identified 117 differentially methylated loci involved in numerous biological processes that may identify cyanogenic- sensitive regions of the genome, providing the first study of epigenomic alterations associated with sub-lethal cyanide exposure and a clinical phenotype.

## Introduction

The World Health Organization (WHO) classifies konzo as a neurological disease characterized by sudden onset of spastic paraparesis in a formerly healthy individual and is strongly correlated with the monotonous consumption of insufficiently processed bitter cassava *(Manihot esculenta Crantz*) and malnutrition, specifically a diet deficient in sulfur amino acids (1,2). Cassava is a fiberous tuber and the predominant food source for many regions throughout in Sub-Saharan Africa, including Cameroon, Central African Republic, the Democratic Republic of the Congo (DRC), Mozambique, and United Republic of Tanzania, which are the only countries in the world where konzo is endemic (1). In addition to konzo, consumption of cyanogenic cassava has also been associated with other complex neurodegenerative syndromes in Sub-Saharan Africa, including tropical ataxic neuropathy and motor neuron-cerebellar-Parkinson-dementia syndrome (1,3,4). Konzo is predominantly linked to rural regions in the five aforementioned countries in Sub-Saharan Africa that lack healthcare infrastructure and resources(1). The mean annual incidence rate of konzo diagnosis is 0.9 per 100,000(1). Studies have determined that cyanide exposure from cassava is associated with a loss of approximately 2 disability adjusted life years and an overall case-fatality ratio of about 21%, making konzo among the most prevalent of any disease associated with chemical exposure through the food supply(1).

Konzo onset typically occurs within one week of cassava consumption in a formerly healthy individual, beginning with bilateral spastic movements of the lower limbs that affect gait and progressing to exaggerated knee and ankle spasms, and occasionally full lower limb paralysis (2,3,5). The disease is non-progressive and irreversible, without any apparent disease to the spinal cord (2,5). Konzo can present in a spectrum of severities and occur with comorbidities, such as cognitive impairment (4,5). Epidemeological surveys have highlighted that children and women of child-bearing age appear to be more vulnerable to developing konzo, and familial clusters of disease occur for reasons that have yet to be fully elucidated(6).

Uncovering the molecular underpinnings of konzo etiology and its comorbidities are of considerable interest, as cassava is a dietary staple for over 800 million people due to its drought tolerance (1). All cultivars of cassava have an innately high concentration of the cyanogenic glucosides, predominantly linamarin, which assist in defending the tuber from animals and insects (1). Proper processing of cassava makes it safe to consume and is crucial to reducing the concentration of linamarin and downstream cyanide exposure (1,4). During times of drought, famine, and war, when resources and time are scarce, proper processing techniques diminish and epidemics of konzo become rampant (1,4).

As konzo can affect up to 10% of the population in regions throughout Sub-Saharan Africa that subsist on a homogenous diet of cassava, it is of critical importance to elucidate the pathophysiologic factors that contribute to disease susceptibility and onset (3). As epigenetics are known to be heavily influenced by the environment, age, and diet, this study focuses on detecting the underlying epigenomic signatures in individuals exposed to a diet of cyanogenic cassava that develop konzo compared to age-matched, unaffected controls from the same outbreak zones in the DRC. DNA methylation has been shown to be a genomic marker of exposure to certain chemicals and thus can be used as a diagnostic signature for poorly understood, environment-induced diseases(7). By highlighting loci within the genome that are differentially methylated we will be able to better understand the individual susceptibility risks of konzo by consumption of cyanogenic cassava, specifically those implicated in the motor phenotype. This is the first study focused on understanding the underlying epigenomic signatures, or biomarkers, associated with sub-lethal cyanide exposure and konzo. Future studies should aim to focus on functional validation of the apparent molecular differences highlighted in this study, including toxicity and adverse outcome pathways, and explore the complex epigenetic and transcriptional landscape associated with sub-lethal cyanide exposure and the clinical konzo phenotype.

## Methods

### Patient Samples

DNA samples (n=32) were obtained from the lab of Desire Tshala-Katumbay MD, PhD, MPH, (Oregon Health & Science University, Portland, Oregon; University of Kinshasa, Kinshasa, DRC) and stored at 4°C until usage. DNA was previously extracted from blood of healthy (n=16) and konzo-affected (n=16) individuals by the Tshala-Katumbay lab at two sites in DRC (Kahemba and Bukavu). Individuals were phenotypically diagnosed using neurological exam performed by neurologists to confirm konzo diagnosis using the 1996 WHO criteria. From a previously established konzo WHO surveillance list of outbreaks in Kahemba and Bukavu, subjects were identified and used in this study (8). For konzo-affected individuals, konzo severity was described as “mildly abnormal (periodic or mild hyper-reflexia and exaggerated clonus or reflexive delay)”(8). Samples from the konzo outbreak regions of Kahemba, southern Bandundu Province, DRC, were obtained in October to November 2011; samples from the konzo outbreak region of Bukavu, South Kivu Province, were collected in November 2012. Patient demographic data can be found in **Supplementary Table 1** and additional details pertaining to the method of diagnosis and procurement of samples can be found in our previous 2013 publication by Boivin *et al*.(8).

### Genome-Wide DNA Methylation Array

DNA was quantified using the Qubit dsDNA Broad Range Assay kit and Qubit 4 fluorometer (Thermo Fisher). 300ng of extracted DNA from each individual was bisulfite converted using the EZ DNA Methylation-Lightning kit (Zymo). Samples were whole-genome amplified, enzymatically fragmented, and hybridized to BeadChip array using the Infinium MethylationEPIC BeadChip kit according to the manufacturer’s protocol (Illumina). Hybridized methylation arrays were scanned using the iScan system (Illumina) at Georgetown University’s Genomics & Epigenomics Shared Resource Core in Washington, DC. These data were deposited for public accessibility in the Gene Expression Omnibus (accession number GSE18011).

### Methylation Array Analysis

Raw intensity values (idat files) were generated from each EPIC BeadChip row using the iScan system. All statistical analysis was done using R (version 4.0.1) and Bioconductor packages. Intensity values were converted to beta (β) and m-values using the Bioconductor *minfi* package. β-values (β=M/(M+U+100), where M is methylated intensity and U is unmethylated intensity for the same position) are used for visualization and m-value (M=log2(M/U)) for statistical analyses. Quality filtration parameters, used to filter out low quality probes were: probes that are represented by less than 3 beads for greater than 5% of the samples (49,632 probes), probes having quality p-value less than 0.05 (15,032 probes), probes having SNP (9,956 probes), and probes mapping to multiple sites in the genome (“cross-reactive probes”; 41,482 probes). Probes filtered for quality p-values and SNP probes were determined by *detectionP* and *dropLociWithSnps* functions of the *minfi* package, cross-reactive probes by *dropXreactiveLoci* function of the *maxprobes* package and bead related filtration by *beadcount* function of the *wateRmelon* package 9. The probe intensity was normalized by Subset-quantile Within Array Normalization (SWAN). A total of 795,169 quality filtered normalized probes were used for subsequent differential methylation analysis using *limma* package in R. Quality visualization was done using *champ*.*QC* function of *ChAMP* package in R, and PCA plots were done using base R package. Tool information and references can be found in **Supplementary Table 2**. Data quality plots can be found in **Supplementary Figure 1**.

### Gene Ontology Enrichment Analysis

Functional annotation was done using Enrichr (https://maayanlab.cloud/Enrichr/) choosing only gene ontology terms enriched for “Biological Processes”. Duplicated or redundant gene ontology terms were reduced through the REVIGO interface using standard parameters and visualized using GOplot (details in Supplementary Table 2).

## Results and Discussion

Comparative analysis of normalized intensities derived from interrogation of over 850,000 methylation probe sites between konzo cases and age-matched healthy controls suggests that there are 117 differentially methylated probes (DMPs) significantly associated with konzo-affected individuals (**Figure 1A**). Of the 117 total sites of differential methylation, 99 DMPs were hypomethylated, while 18 sites were hypermethylated compared to controls (Table S3). Unsupervised hierarchical clustering of DMP intensities revealed that samples cluster strongly by cohort, with the exception of one sample (K1), which appears to cluster more with the control cohort than konzo (**Figure 1A**). Without other clinical information, it is not possible to determine if this sample is clustering differently due to other phenotypic associations (ie. disease onset or severity) or a true outlier of the study.

**Figure 1.**
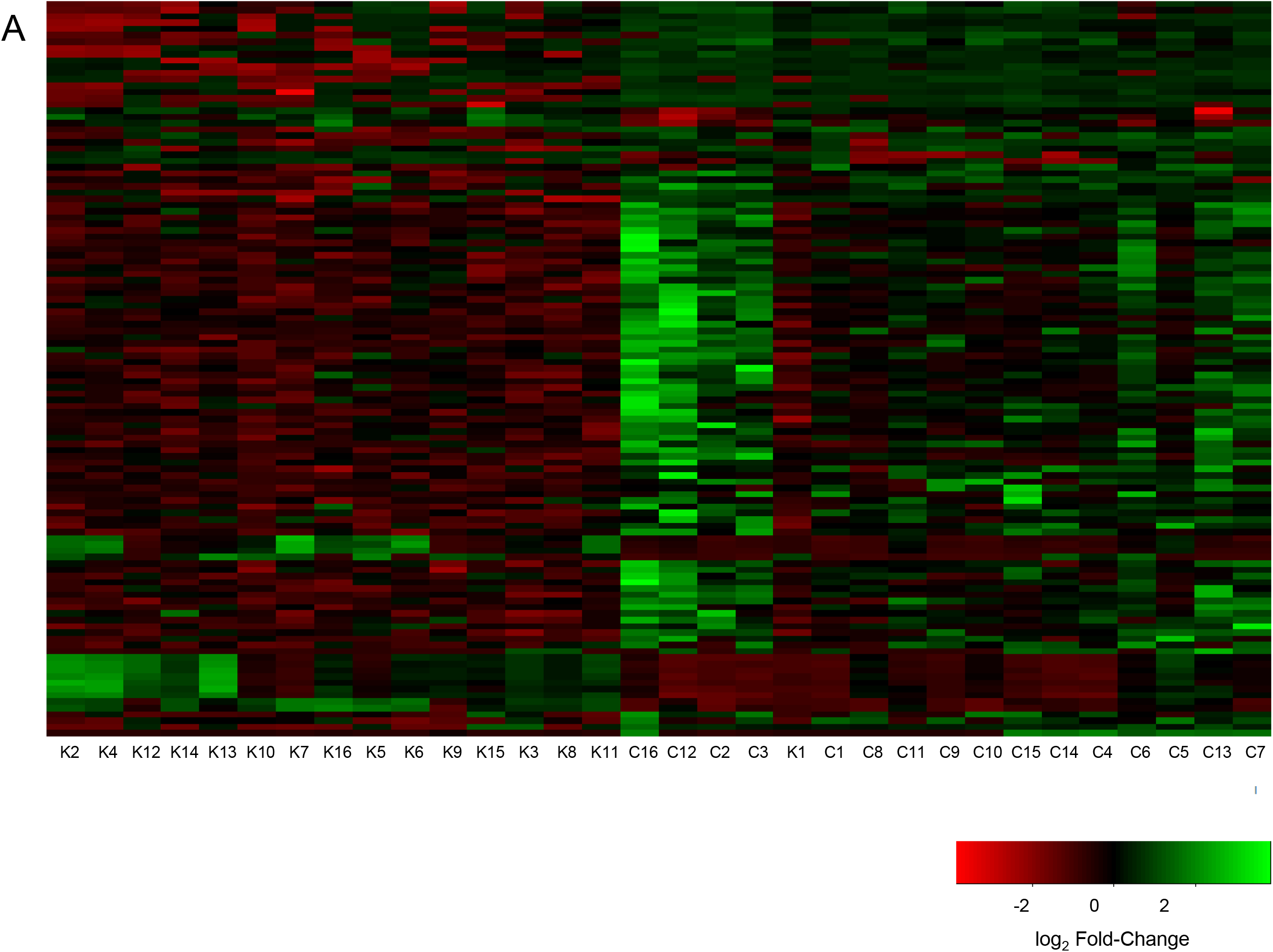
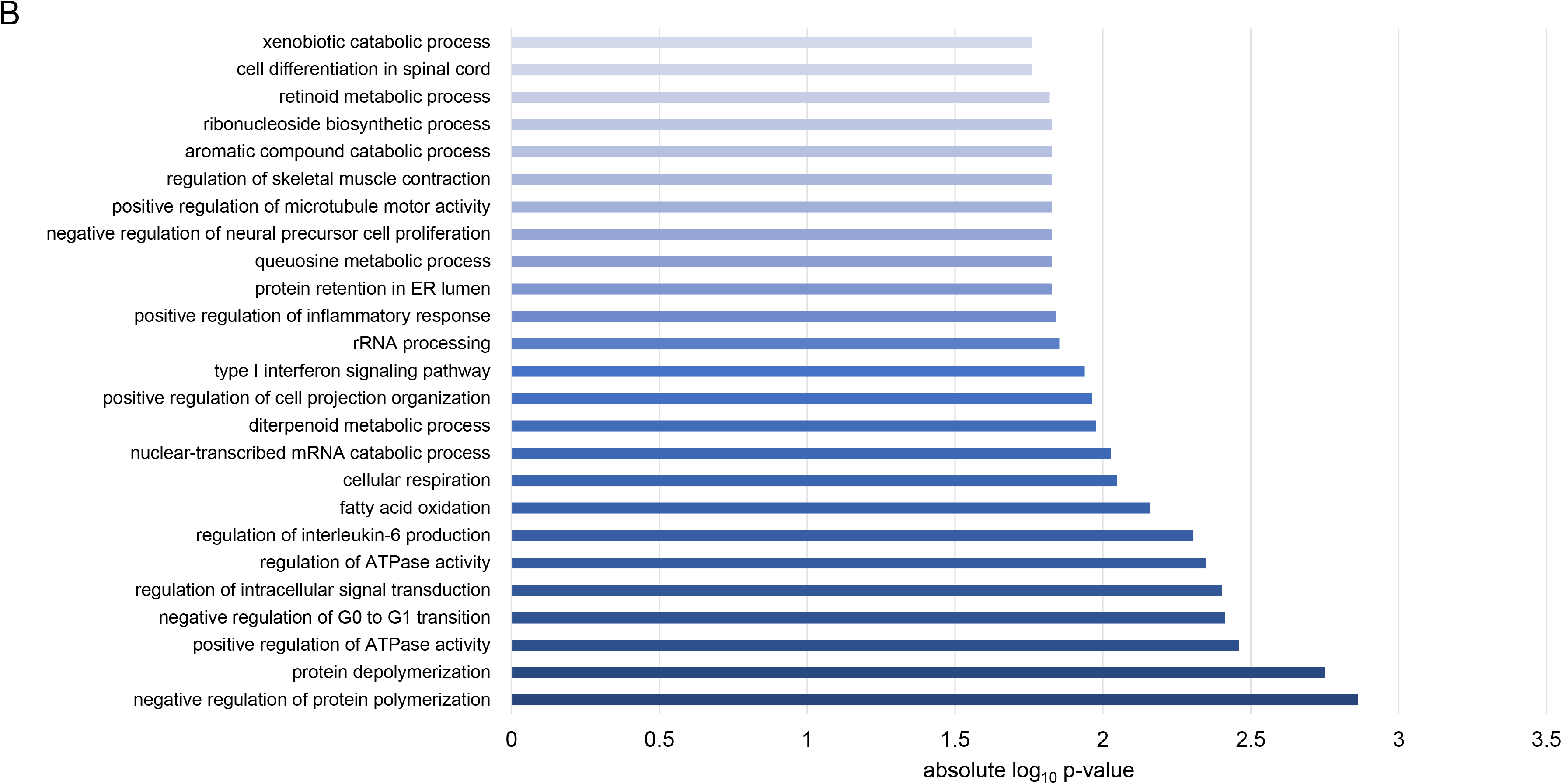
Differentially methylated probes between konzo and control samples reveal unique epigenetic signatures that may relate to clincal presentation and etiology of konzo. A) Heat Map of Unsupervised Hierarchical Clustering of DMPs between konzo cases (K1-16) and controls (C1-16). Limma function in R was used to perform 1-way ANOVA of normalized beta-values between konzo cases and age-matched healthy controls. Significant DMPs were determined using a threshold FDR p-value of < 0.05 and log2 fold-change cut-off of < -1 or > 1. B) Gene Ontology enrichment (GO) analysis of DMPs found in promoter regions (transcription start sites or 5’UTR) associated with konzo. Significant GO terms were determined by an FDR p-value cutoff of < 0.05. Most enriched values were determined using absolute log10 p-value.

Since there are some sex-specific differences in the manifestation and presentation of konzo, where females of child-bearing age appear to be more vulnerable than males (although this may be attributed to social differences), we interrogated the data from a sex difference perspective did not identify significantly differentially methylated loci attributed to sex (data not shown) (5). However, our data also suggests that the analyzed cohort may not have had enough statistical power to uncover sex-specific differences in DNA methylation associated with disease. (5).

Of the 117 DMPs between konzo and controls, there were 2 genes with multiple differentially methylated probes that were significantly differentially methylated in the konzo cohort, *ZNF718* and *AKAP12* (**Supplementary Table 3**). All sites associated with the genes *ZNF718* and *AKAP12* were significantly hypermethylated (FDR p-value ≤ 0.05 and log2 fold-change in DNA methylation intensity ≤1) in konzo cases, compared to controls (**Supplementary Table 3**). 46 of these sites associated with genes were identified in the promoter region (defined as either being located in the transcription start site or 5’ UTR). These 46 sites were analyzed for gene ontology (GO) enrichment and we were able to ascertain that the konzo cohort was enriched for biological processes relevant to konzo etiology and potentially relevant metabolic processes (**Figure 1B, Supplementary Table 4**). For example, among the top 20 most enriched functions, we noted *regulation of skeletal muscle contraction* (GO:0014819), which may be directly relevant to the spastic movements and paraparesis that are characteristically associated with the konzo phenotype (**Figure 1B**)(2). Additionally, using the Online Mendelian Inheritance of Man database, we identified that the associated gene for this GO term, *KCNJ2*, has been implicated in other disorders of periodic paralysis, such as Andersen Syndrome, and thus may be directly relevant to konzo disease presentation (9) (**Supplementary Table 4**).

Of additional interest, we noted significant enrichment for the biological process *queuosine metabolic process* (GO:0046116). Queuosine is a modified nucleoside present in certain mammalian tRNAs and its abundance has been linked to the presence of micronutrients derived from the gut microbiome and directly links to transcriptional regulation (10,11). As konzo onset is linked not only to dietary exposure to cyanogenic glucosides, but also to SAA deficiency, an adjunct role of the gut microbiome could also play into the disease phenotype and be linked to changes in DNA methylation, transcription, and metabolic processes. The literature suggests that DNA methylation is strongly influenced by the environment, so changes in diet that are known to be associated with disruptions in molecular processes and the gut microbiome, like queuosine metabolism, may be of interest for elucidating the complex mechanisms associated with konzo disease onset and progression.

Overall, while enrichment of these biological processes may suggest a role for modifications to DNA methylation in disease phenotype, these 117 sites of differential methylation can serve as biomarkers of monitoring exposure to cyanide in populations at-risk for konzo. Functional validation is critical to further explore these findings and understand the impact of these differentially methylated sites in the context of dietary cyanogenic glucoside exposure and konzo presentation, as well as determine if there are specific epigenetic markers associated with susceptibility or risk to developing a clinical phenotype, as konzo does not present in all who are exposed to the same, homogenous diet of cyanogenic cassava.

A limitation of this study is the small sample size (n=32) that was used. While we were able to determine statistically significant DNA methylation differences between our cohorts, future studies should look to expand on the size of the cohort used, increase the age range, and include numerous disease severity levels, and ensure that a sufficient sample size is used for a well-powered study. Additionally, by correlating the level of *in vivo* cyanogenic glucoside metabolites in serum or urine at the time of collection could provide invaluable information regarding the level exposure of each individual to cyanogenic cassava through the diet and further be associated to the DNA methylation changes present. As previously mentioned, children are also at high-risk groups for developing konzo. In this cohort, the median age of recruited konzo and healthy control individuals was approximately 13 years old. As such, we are unable to draw conclusions regarding the DNA methylation patterns associated with pediatric versus adolescent ages groups. Future research should aim to observe longitudinal progression of this disease and consider timing of onset and severity of these age groups, which may elucidate the molecular underpinnings of the sudden and irreversible phenotype associated with konzo.

This study has provided the first analysis of epigenetic changes associated with konzo and cyanide exposure through the diet. Future experiments should focus on further identifying biomarkers of early low-dose cyanide exposure and identifying factors of konzo disease susceptibility and pathobiology through other molecular approaches.

## Data Availability

The datasets generated and analysed during the current study are available in the GEO repository (GSE180119), https://www.ncbi.nlm.nih.gov/geo/query/acc.cgi?acc=GSE180119. These data will be made publically available on December 31, 2021, and can be made available from the corresponding author prior to this date upon request.

## List of Abbreviations

WHO: World Health Organization
DRC: The Democratic Republic of the Congo
DMP: Differentially methylated probe
SWAN: Subset-quantile within array normalization
GO: Gene ontology

## Declarations

### Ethics Approval and Consent to Participate

Informed consents were obtained verbally by investigators who are fluent in Lingala and/or Kikongo the local spoken languages. Ethical approval of research activities including informed consent was obtained from the Oregon Health & Science University (OSHU) IRB FWA00000161 and from the Ministry of Health of the Democratic Republic of the Congo (DRC).

### Consent for Publication

Not applicable.

## Authors’ Contributions

K .K: quality control and bisulfite conversion of DNA samples, Infinium MethylationEPIC BeadChip array protocol and scanning of arrays, collection of raw data and bioinformatic analysis. S.B.: optimization of bioinformatic pipeline and analysis of data, generation of figures and tables. M.B.: experimental design, data analysis. D.O.L. and D.M.N.: recruited, consented, and collected participants for study in DRC. D.T.K: design and ethics approval for study, supervised data analysis and interpretation. E.V.: experimental design, supervised data analysis and interpretation. All authors contributed to the content of the manuscript.

## Acknowledgements

We would like to thank the Congolese participants of this study for kindly donating specimens for this study. We would also to acknowledge the passing of Jean-Pierre Banea Mayambu, a pioneer in the field of konzo, who will truly be missed.

## Supplemental Figure Legend

**Supplemental Figure 1.**
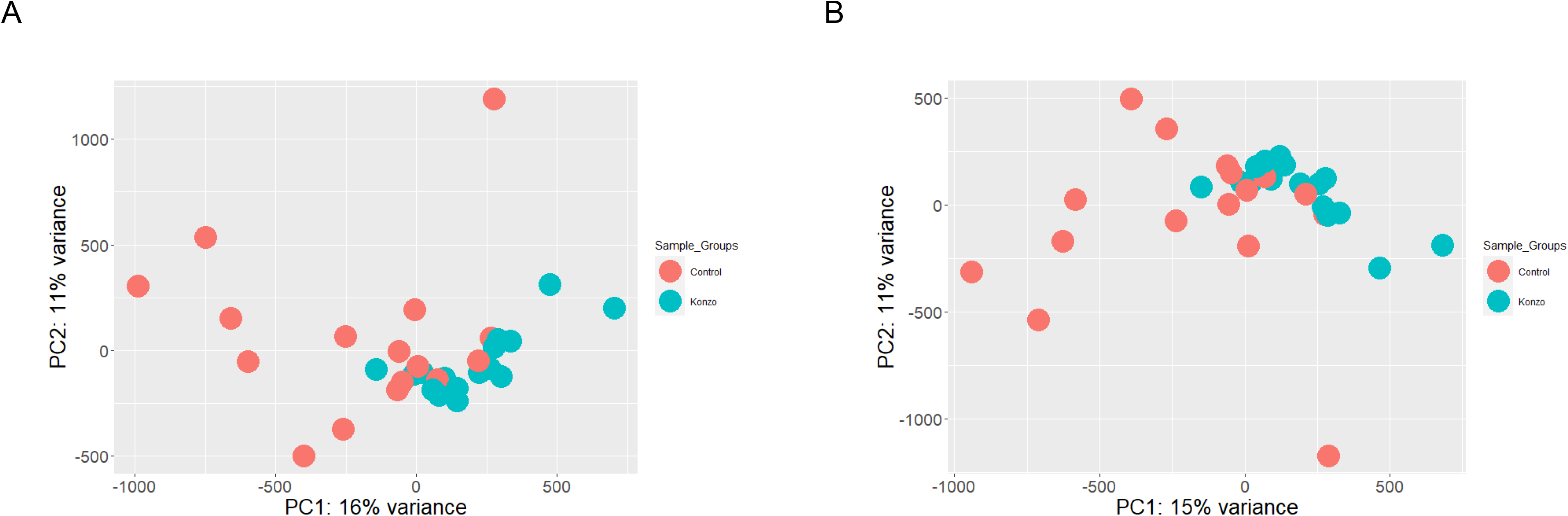
Principal component analysis (PCA) plots shows similarity in signal intensity between konzo and control samples. A) PCA plot for signal intensity of all control and konzo samples before normalization: Principal component analysis done on more than 850 K probes (866,087 probes), reveals a tighter overlap of intensities (principal components) between Konzo samples (blue) compared to red (Control) samples. There is some overlap with between the controls and the Konzo sample, which might lead to the assumption that there is not a huge difference in the epigenetic patterns between konzo and control samples. B) PCA plot for signal intensity of all control and konzo samples after filtration and SWAN normalization: Principal component analysis done on more than 795,169 probes, reveals similar pattern to before normalization, leading to the assumption that there is not much difference in epigenetic signature between the conditions.

## Supplemental Table Legends

**Supplemental Table 1.** Sample collection information. Analysis ID is a deidentified label used for improved readability on figures and to indicate the cohort that each sample belongs to. Cohort specific information, and location of sample collection. The average age of participants is 13 years old.

**Supplemental Table 2.** DNA Methylation analysis tools, functions and references.

**Supplemental Table 3.** Output of significantly differentially methylated probes from bioinformatic analysis. Threshold for significance: FDR p-value < 0.05, log2 fold-change of less than or equal to -1 or greater than or equal to 1. Column headers include the Illumina ID (IlmnID), Chromosome Number (Chromosome) and location (Coordinate_start or Coordinate_end), positive or negative strand (Strand), gene name (Name), location of probe within gene/CpG (Group), log2 fold-change value (logFC), and FDR adjusted p-value (adj.P.Val).

**Supplemental Table 4.** Output of significantly enriched GO Terms. Threshold for significance: p-value < 0.05. Column headers include the GO ID (ID), GO Term Description (GO Description), absolute log10 p-value (abs_log10_pval), log10 p-value (log10_pval), p-value (pval), and genes associated with GO ID (genes).

